# Optimal management of isolated left vertebral artery in total arch replacement with frozen elephant trunk for aortic dissection

**DOI:** 10.1101/2025.02.14.25322315

**Authors:** Sangyu Zhou, Yanxiang Liu, Bowen Zhang, Luchen Wang, Ruojin Zhao, Mingxing Xie, Xuyang Chen, Cuntao Yu, Yaojun Dun, Xiaogang Sun

## Abstract

**Background:** The presence of an isolated left vertebral artery (ILVA) in patients with aortic dissection (AD) is a rare and challenging condition. This study aims to determine the optimal management of ILVA in patients with AD undergoing total arch replacement with frozen elephant trunk (TAR with FET).

**Methods and Results:** This retrospective study enrolled 94 patients with ILVA and AD who underwent TAR with FET. Patients were divided into three groups: 18 patients underwent ligation of ILVA, 52 underwent ILVA- left subclavian artery (LSCA) transposition, and 24 underwent ILVA- left common carotid artery (LCCA) transposition. Vertebral artery dominance was left-dominant in 10.6%, symmetric in 33.0%, and right-dominant in 56.4% of patients. Notably, patients who underwent ligation of ILVA had either symmetric or right-dominant vertebral arteries, with no left-dominant cases. No strokes were observed. Paraplegia/paraparesis, mechanical ventilation time, and long-term survival were comparable among the three groups. Follow-up computed tomographic angiography (CTA) confirmed patency of the left vertebral artery in all patients who underwent ILVA transposition.

**Conclusions:** Ligation of ILVA, ILVA-LSCA transposition, and ILVA-LCCA transposition are all feasible and safe strategies for managing ILVA in patients with AD undergoing TAR with FET. However, ligation of ILVA is not recommended for patients with left-dominant vertebral arteries.

## INTRODUCTION

Aortic dissection (AD) is a life-threatening cardiovascular disease with an annual prevalence of 3-16 cases per 100,000 individuals^1^. For medical management, AD has a mortality rate of 0.5% per hour and 23.7% at 48 hours, but this decreases to 4.4% with surgical management^2^. Total arch replacement with frozen elephant trunk (TAR with FET) has emerged as a major surgical approach for AD involving the aortic arch^3^. This technique has gained favorable results with 10-year survival rate of 81.4%^4^.

An isolated left vertebral artery (ILVA) is the left vertebral artery directly arising from the aortic arch, usually between the left common carotid artery (LCCA) and the left subclavian artery (LSCA). ILVA is a congenital aortic arch anomaly with a prevalence of 2.8% - 3.8%^5–8^. The vertebral arteries terminate in the posterior inferior cerebellar artery. Mismanagement of ILVA during surgery can lead to posterior circulation stroke or spinal cord ischemia, particularly in patients with an incomplete circle of Willis^9^.

The coexistence of ILVA and AD is exceptionally rare and poses significant surgical challenges. More rigorous surgical strategies are needed in patients with ILVA and AD, especially the reconstruction of arch vessels. Regarding management of ILVA, our center have been adopting three approaches including ILVA-LSCA transposition, ILVA-LCCA transposition, and ligation of ILVA. However, there is no consensus on the optimal approach for ILVA management during TAR with FET. Previous studies have been limited by small sample sizes^10–12^. This study reviewed our experience with ILVA management in patients undergoing TAR with FET for AD, aiming to determine the optimal management of ILVA.

## PATIENTS AND METHODS

### Patients

Between January 2015 and February 2024, 116 patients with ILVA and AD who underwent TAR with FET at the Aortic and Vascular Center in Fuwai Hospital were retrospectively enrolled. Exclusion criteria included age <18 years, prior aortic arch replacement, and ILVA combined with other aortic arch anomalies (e.g., bovine arch, aberrant right subclavian artery, or right-sided aortic arch). A total of 94 patients were included in the final analysis. They were divided into three groups: 18 patients underwent ILVA ligation, 52 underwent ILVA-LSCA transposition, and 24 underwent ILVA-LCCA transposition (Figure 1). This study was approved by the ethics committees of Fuwai Hospital (No. 2023-2061). The requirement for informed consent was waived due to the study’s retrospective design.

**Figure 1.**
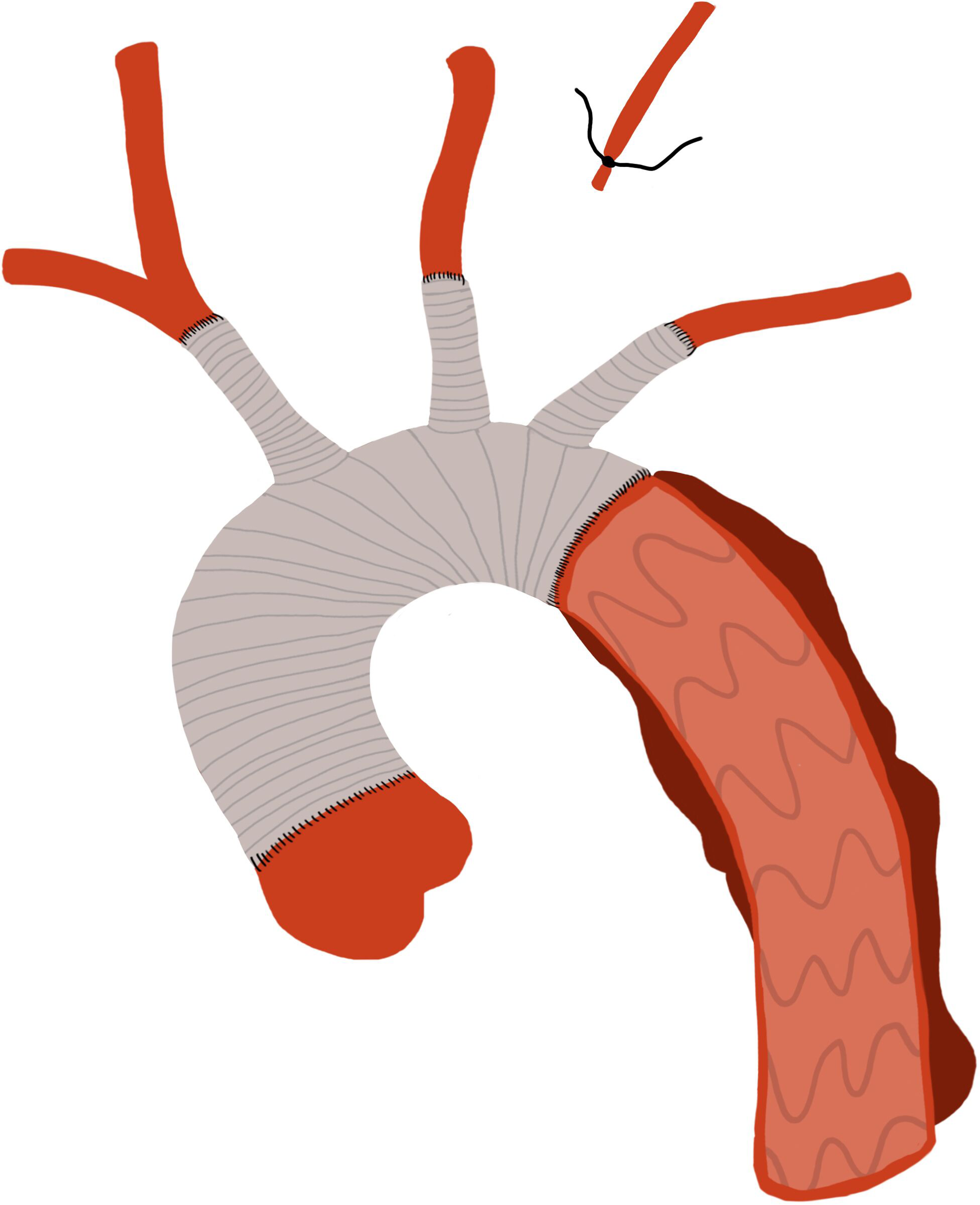
Flow chat.

### Surgical procedure

After median sternotomy, the femoral artery, the right axillary artery, and the right atrium were cannulated for cardiopulmonary bypass. During the cooling phase, the operation on the aortic root and ascending aorta was performed as indicated. When the nasopharyngeal temperature reached the target temperature, the lower-body circulatory arrest began with selective cerebral perfusion. Unilateral cerebral perfusion was through the right axillary artery, while bilateral cerebral perfusion was through the right axillary artery and LCCA. After the aortic arch was transected between the LCCA and the LSCA, a FET stent graft (Cronus, MicroPort Endovascular Shanghai Co, Ltd, China) was inserted into the descending aorta. Subsequently, with its proximal end being clamped, the distal end of a tetrafurcate graft (Terumo, Vascutek Limited, Renfrewshire, UK) was anastomosed to the descending aorta and the FET together. Once the distal arch anastomosis completed, the lower-body circulatory was restored through the perfusion limb of the tetrafurcate graft. Afterwards, the LCCA was anastomosed to one limb of the tetrafurcate graft, and the rewarming phase began. The proximal end of the tetrafurcate graft was anastomosed to the ascending aorta or repaired aortic root, and the innominate artery and the LSCA were anastomosed to the other two limbs, respectively.

Regarding the management of ILVA, ILVA-LSCA transposition or ILVA-LSCA transposition was performed after the transposed artery was anastomosed to the tetrafucate graft (Figure 2A&2B). Ligation of ILVA was performed before the transection of the aortic arch (Figure 2C).

**Figure 2.**
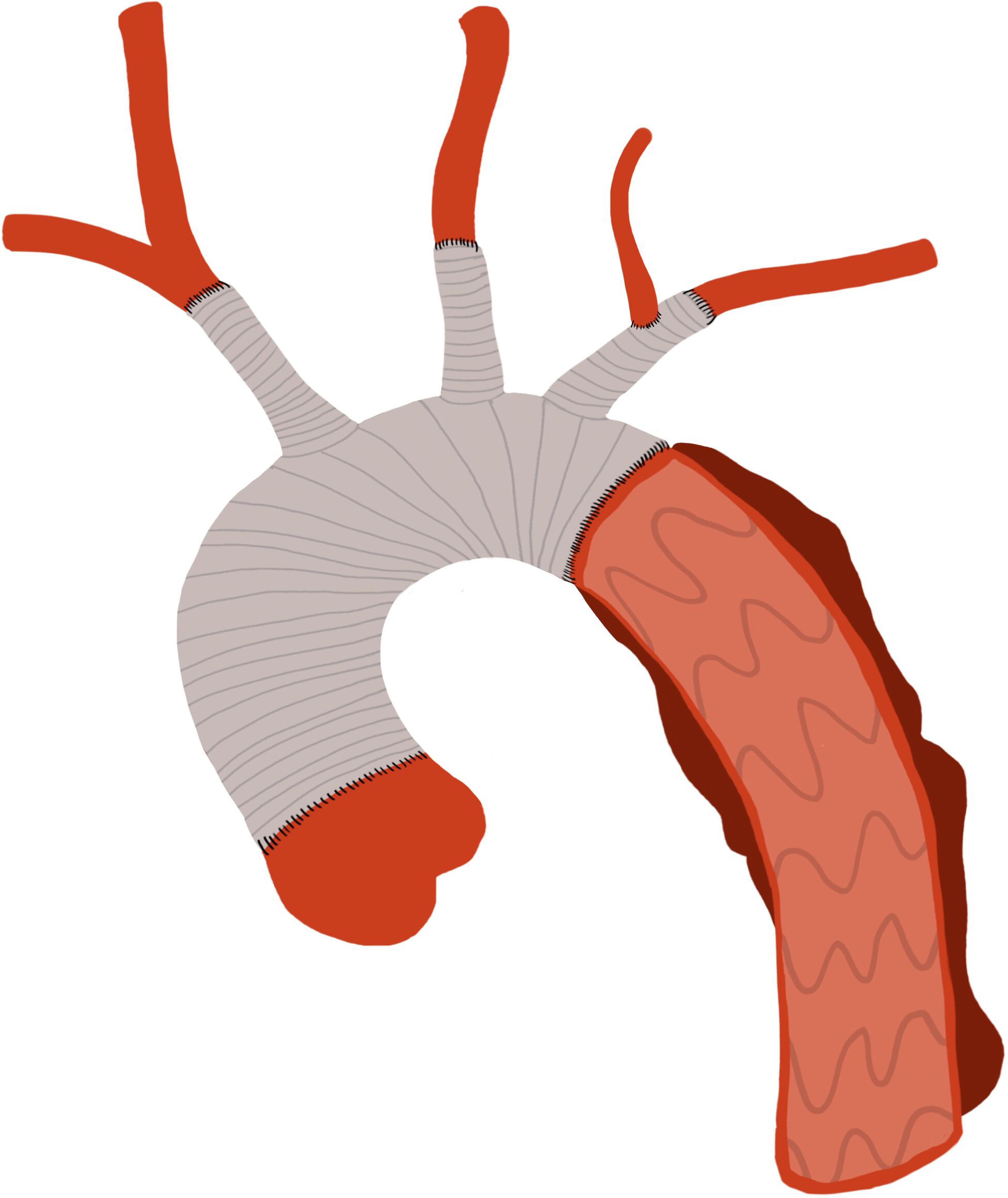
Management of ILVA. **(A)** Ligation of ILVA. **(B)** ILVA-LSCA transposition. **(C)** ILVA-LCCA transposition. ILVA, isolated left vertebral artery; LSCA, left subclavian artery; LCCA, left common carotid artery.

### Data definition and follow-up

Baseline characteristics, imaging findings, intraoperative details, and outcomes were retrospectively collected. Computed tomographic angiography (CTA) was preoperatively performed to confirm the presence of ILVA, its dominance, and associated findings. Left ventricular ejection fraction, aortic regurgitation, and abnormal ventricular wall motion were acquired from echocardiographic reports.

Malperfusion syndrome was diagnosed based on clinical symptoms (coma, altered mental status, abdominal pain, sensory or motor dysfunction of extremity), laboratory tests (elevated troponin, liver enzymes, serum creatine, lactate), and imaging evidence (CTA). Low cardiac output syndrome was defined as requiring intra-aortic balloon pump or extracorporeal membrane oxygenation support. Paraplegia/paraparesis was defined as complete or partial loss of lower limb motor function related to spinal cord ischemia and not related to stroke until hospital discharge.

Follow-up included outpatient visits and telephone consultations. All patients had an aortic CTA at discharge and were recommended to have an aortic CTA at 3 months and annually after surgery.

### Statistics analysis

Continuous variables were expressed as mean ± standard deviation or median (the first interquartile, the third interquartile) and assessed with Student’s t-test or Mann-Whitney U test. Categorical variables were expressed as count (frequency) and compared by Chi-squared or Fisher’s exact test. Kaplan-Meier analysis was performed to analyze the overall survival among three groups. All statistical tests were 2-tailed and P<0.05 was considered statistically significant. Statistical analysis was assessed using R version 4.4.2 (The R Foundation for Statistical Computing, Vienna, Austria).

## RESULTS

### Baseline characteristics

Baseline characteristics are presented in Table 1. Acute Type A aortic dissection was present in 88.9%, 84.6%, and 75.0% of patients for ligation of ILVA, ILVA-LSCA transposition, and ILVA-LCCA transposition, respectively. One-fifth to one-third of patients had at least one kind of malperfusion syndrome. Vertebral artery dominance was left-dominant in 10.6%, symmetric in 33.0%, and right-dominant in 56.4% of patients. Notably, no patients with left-dominant vertebral arteries underwent ligation of ILVA.

**Table 1.**
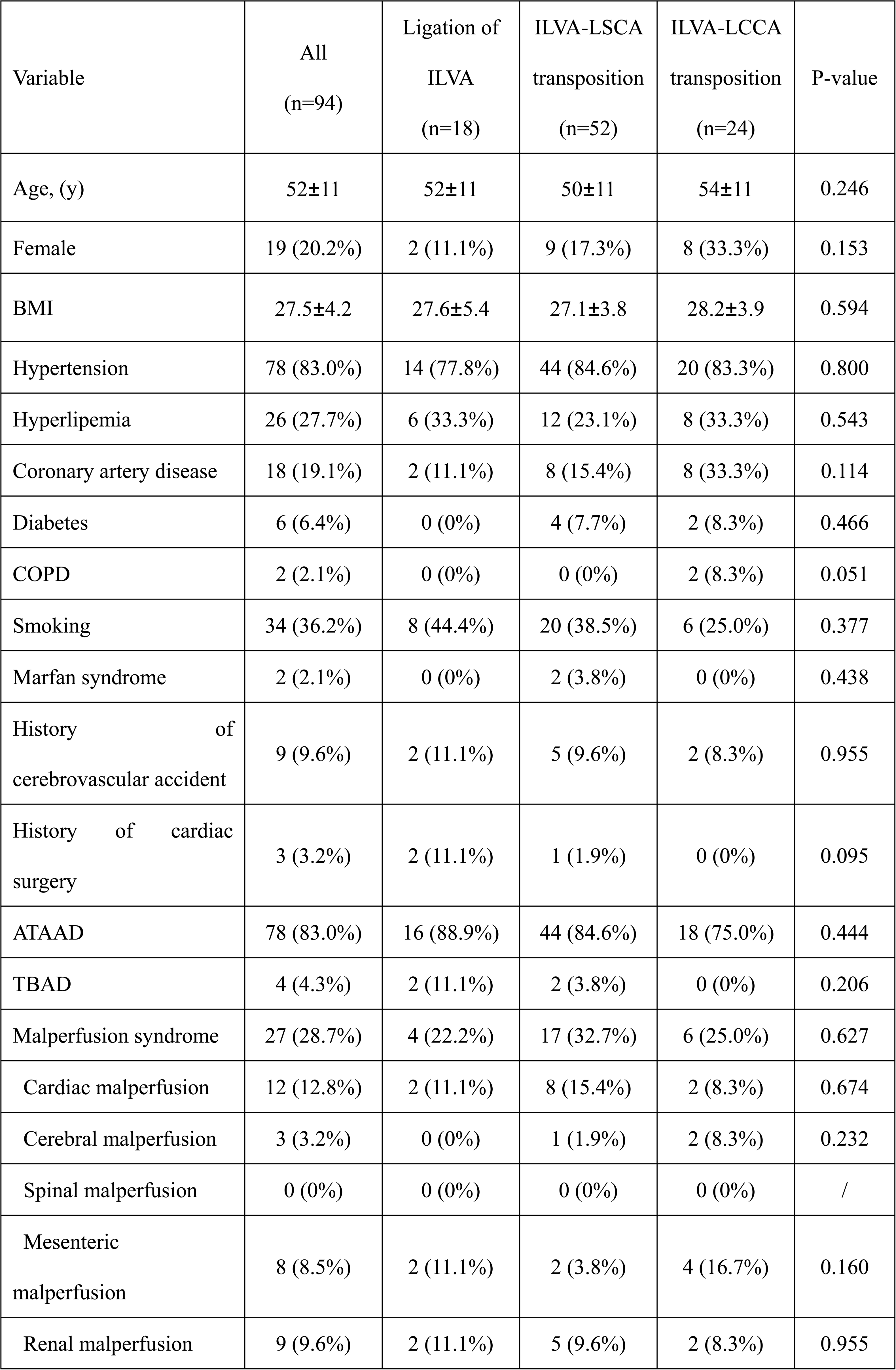
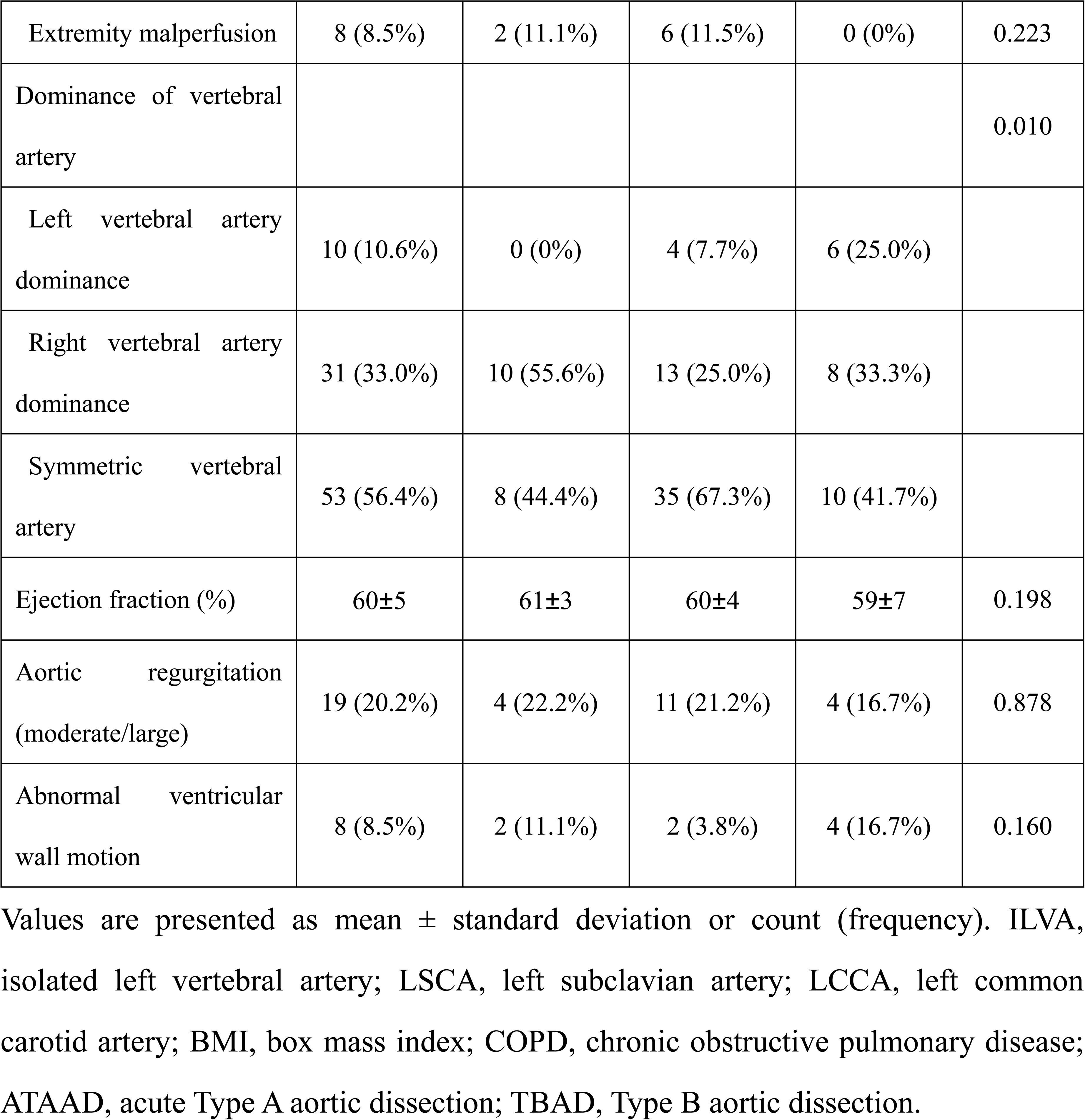
Baseline characteristics.

### Intraoperative data and postoperative outcomes

Intraoperative data are shown in Table 2. Almost half of the patients had unilateral antegrade cerebral perfusion, and the other had bilateral antegrade cerebral perfusion both in the total cohort and each group. Concomitant procedures were comparable among groups. Patients who underwent ligation of ILVA had shorter cardiopulmonary bypass time [147 (134, 191) min vs 187 (165, 211) min for ILVA-LSCA transposition vs 214 (157, 260) min for ILVA-LCCA transposition, P=0.021] and cross-clamp time [87 (67, 115) min vs 117 (98, 137) min for ILVA-LSCA transposition vs 122 (88, 171) min for ILVA-LCCA transposition, P=0.002].

**Table 2.** Intraoperative data.

| Variable | All<br>(n=94) | Ligation of<br>ILVA<br>(n=18) | ILVA-LSCA<br>transposition<br>(n=52) | ILVA-LCCA<br>transposition<br>(n=24) | P-value |
| --- | --- | --- | --- | --- | --- |
| Bentall procedure | 8 (8.5%) | 0 (0%) | 6 (11.5%) | 2 (8.3%) | 0.319 |
| David procedure | 3 (3.2%) | 0 (0%) | 3 (5.8%) | 0 (0%) | 0.286 |
| Concomitant CABG | 17 (18.1%) | 2 (11.1%) | 9 (17.3%) | 6 (25.0%) | 0.021 |
| Cerebral perfusion |  |  |  |  | 0.889 |
| Unilateral | 51 (54.3%) | 10 (55.6%) | 29 (55.8%) | 12 (50.0%) |  |
| Bilateral | 43 (45.7%) | 8 (44.4%) | 23 (44.2%) | 12 (50.0%) |  |
| Lowest nasopharyngeal<br>temperature (°C) | 25.2 (23.1,<br>26.8) | 23.1 (20.1,<br>26.7) | 25.3 (23.9,<br>26.9) | 25.7 (24.9,<br>26.7) | 0.889 |
| Cardiopulmonary bypass<br>time (min) | 185 (149,<br>216) | 147 (134,<br>191) | 187 (165,<br>211) | 214 (157,<br>260) | 0.021 |
| Cross-clamp time (min) | 116 (88,<br>137) | 87 (67, 115) | 117 (98,<br>137) | 122 (88,<br>171) | 0.002 |

Values are presented as mean ± standard deviation, median (the first quartile, the third quartile), or count (frequency). ILVA, isolated left vertebral artery; LSCA, left subclavian artery; LCCA, left common carotid artery; CABG, coronary artery bypass grafting.

Postoperative outcomes are presented in Table 3. Thirty-day mortality was 11.1% (2/18), 5.8% (3/52), and 8.3% (2/24) for patients receiving ligation of ILVA, ILVA-LSCA transposition, and ILVA-LCCA transposition, respectively (P=0.744). No patient had stroke. Paraplegia/paraparesis was observed in 11.1% (2/18), 11.5% (6/52), and 0% of patients for ligation of ILVA, ILVA-LSCA transposition, and ILVA-LCCA transposition (P=0.223). Mechanical ventilation time, ICU stay and hospital stay were comparable among groups.

**Table 3.** Postoperative outcomes.

| Variable | All<br>(n=94) | Ligation of<br>ILVA<br>(n=18) | ILVA-LSCA<br>transposition<br>(n=52) | ILVA-LCCA<br>transposition<br>(n=24) | P-value |
| --- | --- | --- | --- | --- | --- |
| 30-day mortality | 7 (7.4%) | 2 (11.1%) | 3 (5.8%) | 2 (8.3%) | 0.744 |
| Stroke | 0 (0%) | 0 (0%) | 0 (0%) | 0 (0%) | / |
| Paraplegia/paraparesis | 8 (8.5%) | 2 (11.1%) | 6 (11.5%) | 0 (0%) | 0.223 |
| Re-exploration for<br>bleeding | 1 (1.1%) | 0 (0%) | 1 (1.9%) | 0 (0%) | 0.665 |
| Continuous renal<br>replacement therapy | 12 (12.8%) | 4 (22.2%) | 4 (7.7%) | 4 (16.7%) | 0.226 |
| Low cardiac output<br>syndrome | 0 (0%) | 0 (0%) | 0 (0%) | 0 (0%) | / |
| Mechanical ventilation<br>time (h) | 16 (11, 54) | 45 (10, 61) | 18 (11, 38) | 15 (11, 51) | 0.855 |
| ICU stay (d) | 3 (2, 5) | 5 (1, 10) | 3 (2, 5) | 3 (2, 6) | 0.861 |
| hospital stay (d) | 10 (9, 14) | 12 (9, 14) | 10 (9, 14) | 11 (9, 14) | 0.719 |

Values are presented as median (the first quartile, the third quartile), or count (frequency). ICU, intensive care unit.

### Follow-up period

The median follow-up period was 2.6 (1.3, 5.6) years with a follow-up completeness of 96.8%. Three patients were lost to follow-up. Six late deaths occurred during follow-up. Kaplan-Meier curves showed no significant difference among the three groups (log-rank P=0.419) (Figure 3). 1-year and 3-year survival rates were 88.9% and 77.9% for patients with ligation of ILVA, 92.1% and 89.6% for patients with ILVA-LSCA transposition, and 90.9% and 77.9% for patients with ILVA-LCCA transposition, respectively. Three patients underwent a 2-stage thoracoabdominal aortic repair for unsuccessful distal aortic remodeling.

**Figure 3.**
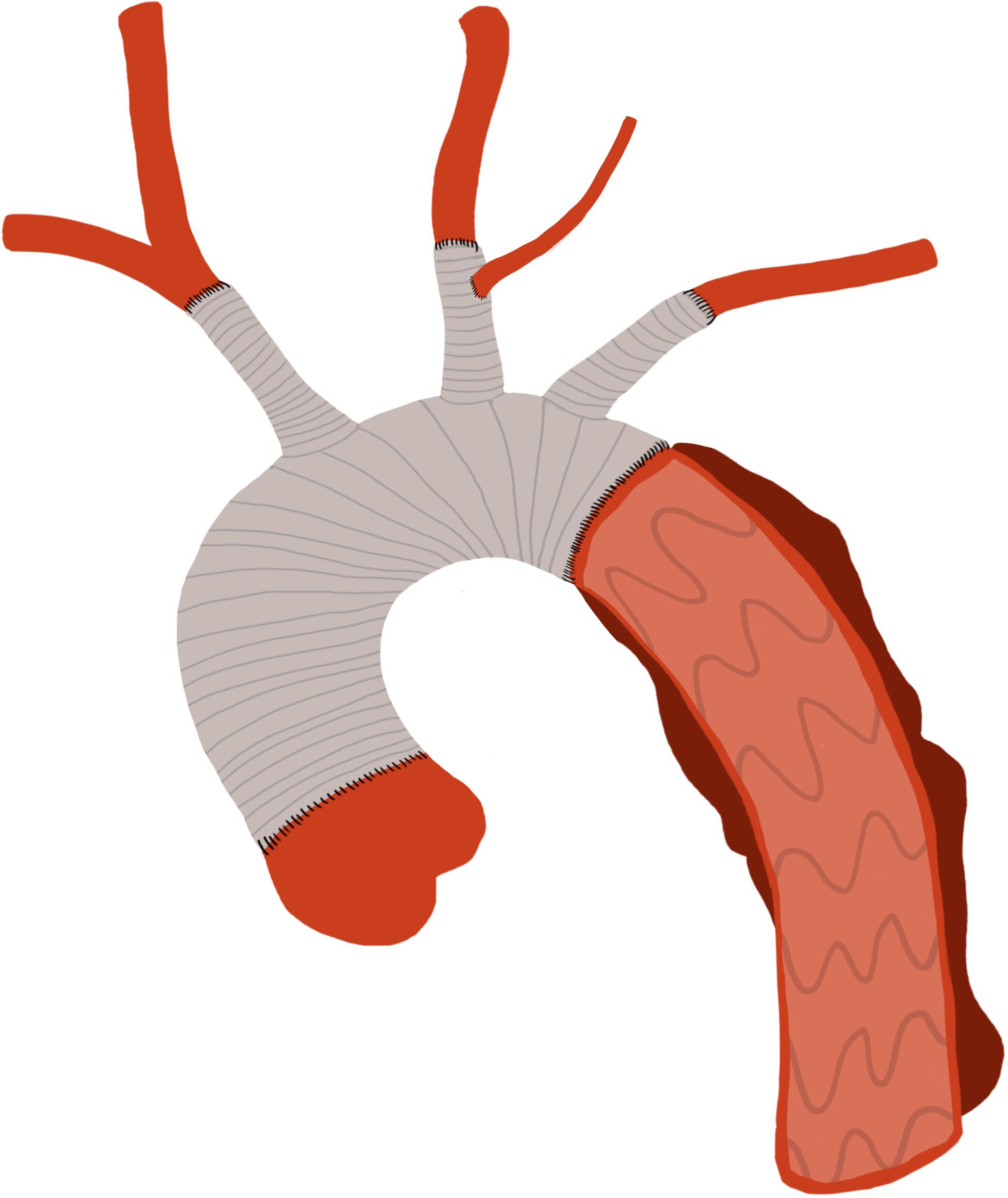
Kaplan–Meier analysis estimating overall survival among patients receiving ligation of ILVA, ILVA-LSCA transposition, and ILVA-LCCA transposition.

Postoperative or follow-up CTA was performed on 85.1% of patients (80/94) at our center with a median period of 4.4 (0.6, 20.2) months. Occlusion of the left vertebral artery was observed in 15.0% (12/80) of patients, all of whom received ligation of ILVA (Figure 4A). In contrast, all patients with transposed ILVA retained a patent left vertebral artery (Figure 4B&4C).

**Figure 4.**
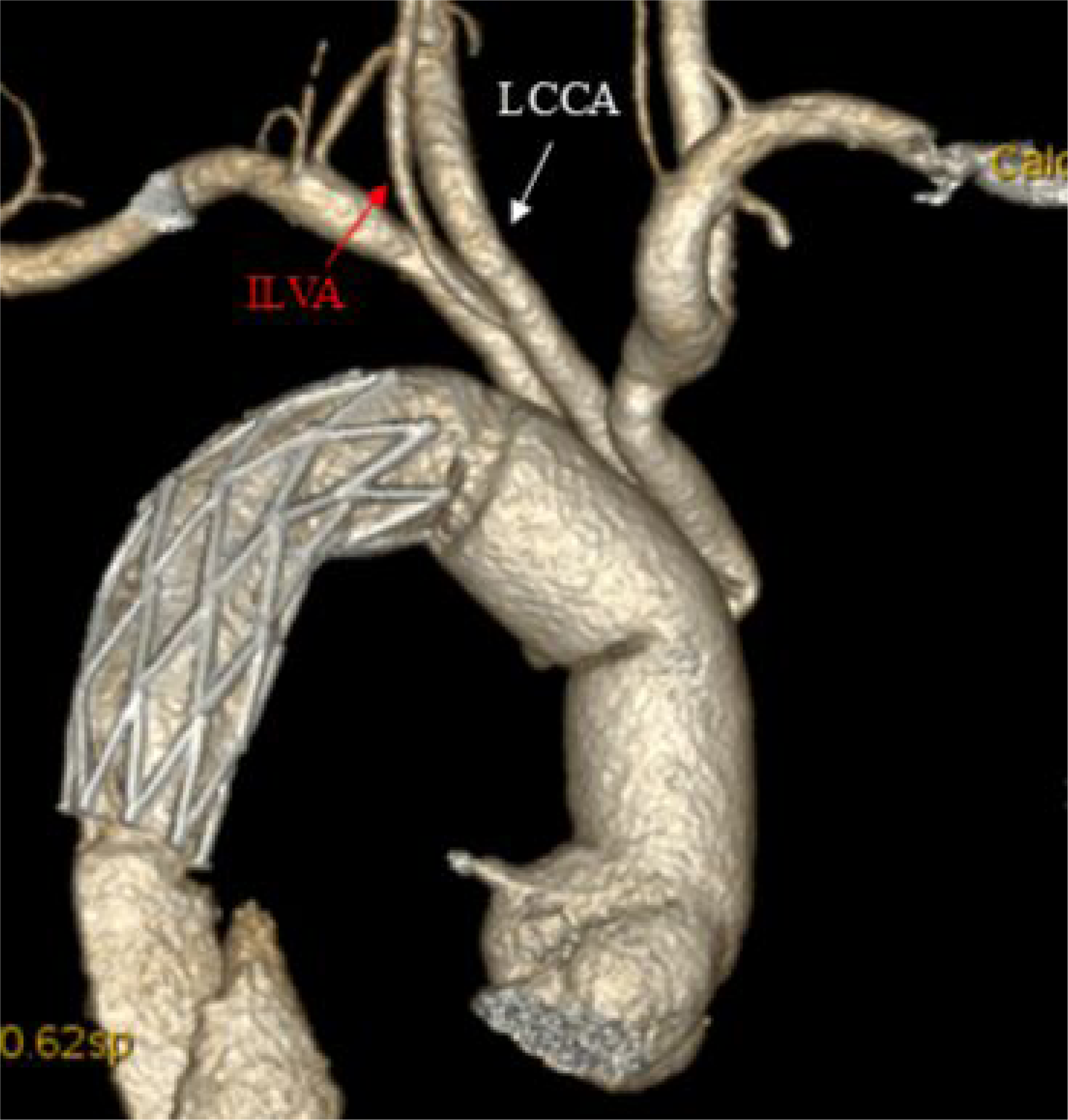
(A) Follow-up CTA of a patient undergoing ligation of ILVA. **(B)** Follow-up CTA of a patient undergoing ILVA-LSCA transposition. **(C)** Follow-up CTA of a patient undergoing ILVA-LCCA transposition. CTA, computed tomographic angiography; ILVA, isolated left vertebral artery; LSCA, left subclavian artery; LCCA, left common carotid artery.

## DISCUSSION

This study found that half of the patients with AD had symmetric vertebral arteries, while only 10% of patients with AD had left-dominant vertebral arteries. Ligation of ILVA was performed only on patients with either symmetric or right-dominant vertebral artery. Patients receiving ligation of ILVA underwent shorter cardiopulmonary bypass time and cross-clamp time. Thirty-day mortality, paraplegia/paraparesis, mechanical ventilation time, and long-term survival were comparable among three groups. No patients had stroke. All patients who underwent ILVA transposition showed patent left vertebral artery in follow-up CTA.

Management of ILVA varied among centers. Suzuki et al performed ILVA-LSCA transposition in TAR for 7 patients and found no neurologic complication^13^. Zhu et al anastomosed the ILVA to the LSCA or the LSCA graft branch during the aortic arch replacement of 22 patients with Type A AD, and 5% of patients had stroke or delirum^14^. Qi et al chose either ILVA-LSCA transposition or ILVA-LCCA transposition in TAR on 21 patients with ILVA and AD^12^. Ten percent of patients had spinal cord injury but recovered in the follow-up period, and 5% of patients had transient neurologic deficit and recovered before discharged. Moreover, there were some novel managements of ILVA. Zuo et al performed prior ILVA-LCCA transposition under normothermic off-pump conditions in TAR on 13 patients with ILVA and Type A AD^10^. They decreased the time spent on treating ILVA during circulatory arrest and avoided using lesion artery wall in the reconstruction of ILVA. No neurologic complications were observed. Zhu et al used the stented elephant trunk technique in 7 patients with ILVA and Type B AD^11^. This strategy was more of an alternative for patients without a suitable proximal landing zone for thoracic endovascular aortic repair. In our center, we performed 3 approaches regarding management of ILVA, including ligation of ILVA, ILVA-LSCA transposition, and ILVA-LCCA transposition. We found that these 3 approaches were not associated with adverse events including stroke, paraplegia/paraparesis, mechanical ventilation time, and long-term survival. Hence, ligation of ILVA, ILVA-LSCA transposition, and ILVA-LCCA transposition were all available in TAR with FET for patients with AD and ILVA.

Regarding the indications for each management of ILVA, firstly, ligation of ILVA was associated with its diameter. Thin ILVA which belonged to either right-dominant vertebral arteries or symmetric vertebral arteries was considered for ligation, after confirming a complete circle of Willis. Ten percent of patients in our cohort had left vertebral artery dominance, consistent with prior research on ILVA and Type B AD^15^. These patients only underwent ILVA-LSCA transposition or ILVA-LCCA transposition. Secondly, the situation of LSCA and LCCA had an impact on the management of ILVA. Transposition was not considered if LSCA or LCCA was involved by AD. Thirdly, the anatomical locations of the arch vessels affected the strategy. Since ILVA was located between LSCA and LCCA, a shorter distance between transposed artery and ILVA was preferable.

Patients with transposed ILVA in our cohort successfully preserved long-term vessel patency of ILVA. Vertebral arteries which converged into the basilar arteries were the primary blood supply for posterior fossa brain structures including the cerebellum and occipital lobe of the cerebrum^16^. As Chinese had a lower incidence of a complete circle of Willis, Ligation of ILVA would increase the risk of postoperative stroke or spinal cord injury^17^. However, these three approaches of management had similar results. It might be that patients receiving ligation of ILVA underwent shorter cardiopulmonary bypass time. Previous research indicated that prolonged cardiopulmonary bypass time was associated with postoperative stroke in TAR with FET^18^. In addition, since ligation of ILVA was performed only on patients with either symmetric or right-dominant vertebral artery, their right vertebral artery and the complete circle of Willis were enough to compensate for the absence of ILVA.

### Limitations

This was a retrospective study from a single-center experience, which might introduce selection bias and limit generalizability. Although the follow-up rate was high, the relatively short median follow-up period may not fully capture long-term complications and survival trends. Future multicenter, prospective studies with longer follow-up durations are needed to validate these results.

## CONCLUSIONS

Ligation of ILVA, ILVA-LSCA transposition, and ILVA-LCCA transposition are all feasible and safe strategies for managing ILVA in patients with AD undergoing TAR with FET. However, ligation of ILVA should be avoided in patients with left-dominant vertebral arteries. These findings provide valuable guidance for surgical decision-making in this rare and complex patient population.

## Data Availability

Data used in this study could be reached from the corresponding authors at reasonable requests.

